# Angiotensin converting enzyme (ACE) 1 expression in leukocytes of adults from 64 to 67 years old

**DOI:** 10.1101/2022.07.27.22278062

**Authors:** Valquiria Bueno, Pedro Henrique Destro, Daniela Teixeira, Daniela Frasca

## Abstract

The renin angiotensin system (RAS) is composed of several enzymes and substrates on which angiotensin converting enzyme (ACE) 1 and renin act to produce angiotensin II. ACE1 and its substrates control blood pressure, affect cardiovascular and renal function, hematopoiesis, reproduction, and immunity. The increased expression of ACE1 has been observed in human monocytes during congestive heart failure and abdominal aortic aneurysm. Moreover, T lymphocytes from hypertensive patients presented increased expression of ACE1 gene after in vitro stimulation with Angiotensin-II (AngII) with the highest ACE1 expression observed in hypertensive patients with low-grade inflammation. Our group and others have shown that aging is associated with comorbidities, chronic inflammation and immunosenescence, but there is a lack of data about ACE1 expression on immune cells during the aging process. Therefore, our aim was to evaluate the levels of ACE1 expression in non-lymphoid as compared to lymphoid cells in association with the immunosenescence profile in adults older than 60 years. Cryopreserved peripheral blood mononuclear cells obtained from blood samples were used. Cells were stained with monoclonal antibodies and evaluated by flow cytometry. We found that ACE1 was expressed in 56.9% of non-lymphocytes and in more than 90% of lymphocytes (all phenotypes). All donors exhibited characteristics of immunosenescence, as evaluated by low frequencies of Naive CD4^+^ and CD8^+^ T cells, high frequencies of EMRA CD8^+^ T cells, and double-negative memory B cells. These findings in addition to the increased C-reactive protein levels are intriguing questions for the study of ACE1, inflammaging, immunosenescence and perspectives for drug development or repurposing.

## Introduction

Angiotensin converting enzyme (ACE1/CD143) and renin are components of the renin angiotensin system (RAS) acting to produce angiotensin II. In a simplistic definition, RAS is composed by a vasoconstrictor, pro-inflammatory ACE1/AngII/AT_1_R axis, and a vasodilating anti-inflammatory ACE2/Ang1-7/MasR axis (Figure 1). In addition to the blood pressure control, ACE1 and its peptide substrates affect cardiovascular and renal function, hematopoiesis, reproduction, and the immunity. [1, 2] Thus, it seems crucial that RAS presents an inflammatory and an anti-inflammatory axis for the adequate regulation of the immune response. ACE1 expression has been observed not only in tissues since its soluble form was found in urine, serum, seminal fluid, amniotic fluid, and cerebrospinal fluid. [3]

**Figure 1.**
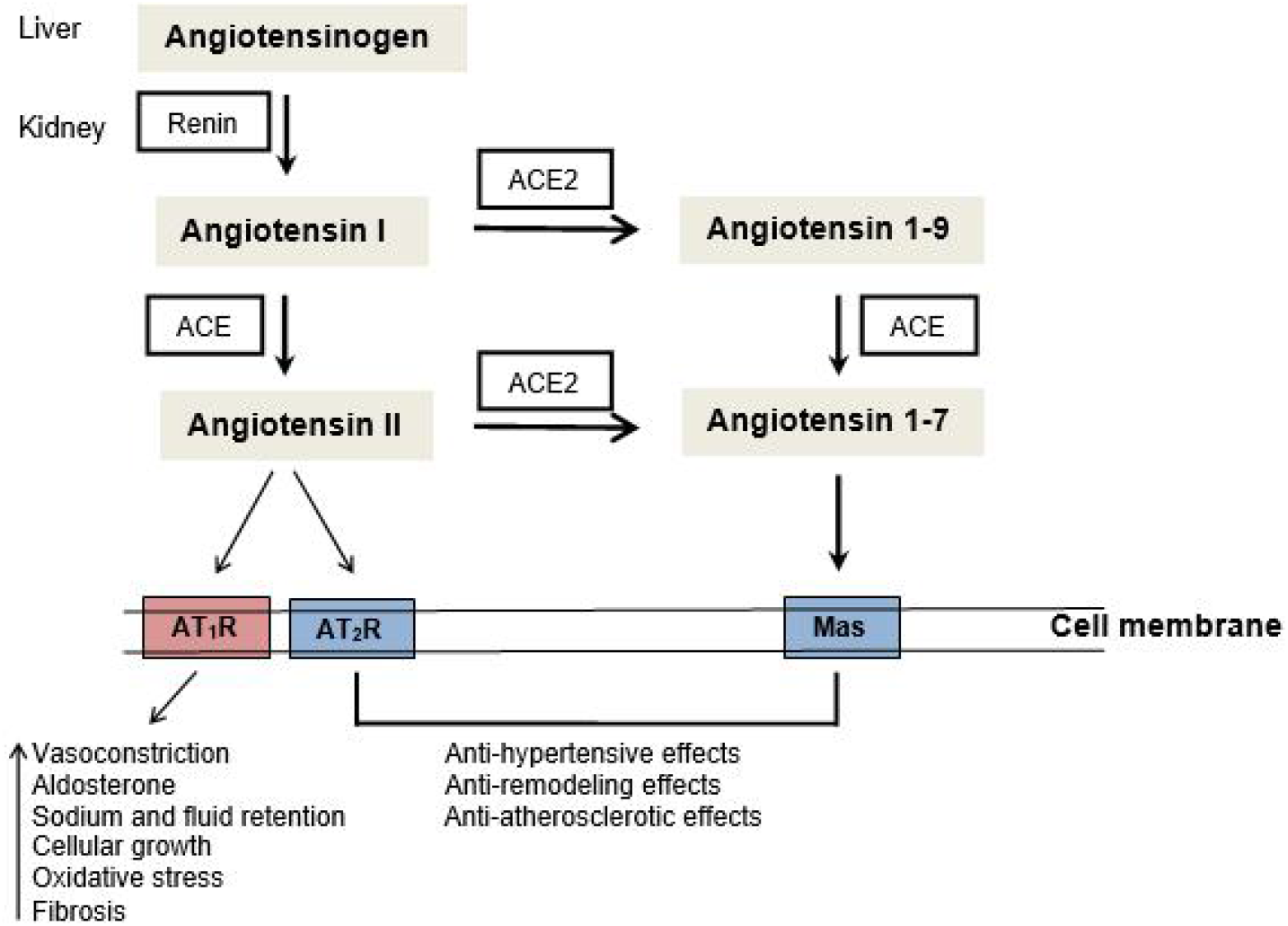
The renin angiotensin system (RAS): ACE1 - angiotensin converting enzyme 1, AT1R - Angiotensin II type 1 receptor, AT2R - Angiotensin II type 2 receptor

The expression of ACE1 in cells from the immune system has been reported in health and disease. Costerousse et al. observed in healthy adult donors by RT-PCR and southern blot the expression of ACE1 in monocytes, macrophages, and T cells but not in B cells. In addition, ACE1 activity was very low in monocytes whereas it was high in macrophages (monocytes driven to differentiation). T cells presented intermediary ACE1 activity and B cells expressed no activity. [4] In patients with type 1 diabetes (29 years, normotensive) it was observed higher ACE1 and lower ACE2 gene expression when compared to healthy (32 years, normotensive) controls. [5] Coppo et al. [6] found that T cells in culture expressed increased mRNA for ACE1 and AT_1_R in obese individuals with low-grade inflammation (high sensitivity C-reactive protein > 3mg/dL). ACE1 activity in the supernatant of T-cell culture was also increased in obese individuals with hsCRP > 3mg/dL. Moreover, RAS gene expression on T cells and levels of inflammatory cytokines in the serum were oppositely associated with serum levels of insulin. [6, 7] Ulrich et al. [8] have shown that the increased expression of ACE1 in monocytes was associated with kidney and cardiovascular disease progression suggesting that circulating leukocytes can modulate local immune response via their own RAS components. [8, 9, 10]

Considering that aging has been associated with comorbidities, low-grade of chronic inflammation and altered frequency/function of immune system cells [11, 12, 13, 14], it seems reasonable to suggest that ACE1 play an important role in the aging process. ACE1 has been suggested to influence age-related diseases (i.e. Alzheimer’s, sarcopenia, cancer) but the associated mechanisms are still under investigation. ACE1 polymorphisms were correlated with susceptibility to Alzheimer’s disease (AD). [15, 16] In addition, it was shown recently that in normal aging ACE1 expression is increased in brain homogenates and this expression is unchanged in early stages of AD. [17] Regarding sarcopenia, Yoshihara et al. [18] found a weak correlation between ACE polymorphism and physical function. In cancer (gastric or colorectal), patients presented higher expression of ACE1 in tumor when compared with healthy tissues. [19, 20] In hematopoietic stem progenitor cells isolated from peripheral blood, Joshi et al. [21] showed that aging is associated with decreased ACE2 and increased ACE1 protein expression. This imbalance suggests a bias to the detrimental pro-inflammatory axis of local RAS. Considering the scarce information about ACE1 expression in the phenotypes of T and B cells, we aimed to investigate ACE1 expression in cells from the immune system and parameters of immunosenescence in adults older than 60 years. Results herein show different levels of expression of ACE1 in non-lymphoid versus lymphoid cells, with the expression being higher in lymphoid cells.

## Materials and Methods

The Ethics Committee of the Federal University of São Paulo - UNIFESP approved all procedures (Protocol number 10904). Blood was collected from adults (n=6, four females and two males) aged 64-67 years old in 2015. Peripheral blood mononuclear cells (PBMCs) were isolated using Ficoll–Hypaque density gradient (Amersham Biosciences, Uppsala, Sweden) and centrifugation. Viable cells were counted, adjusted for 2 × 10^6^/100 μL in 80% fetal bovine serum and 20% dimethylsulfoxide (Sigma, St. Louis, MO, USA), and frozen stored until the phenotyping. In 2021 cells were thawed, checked for viability, and stained with monoclonal antibodies to T-cell phenotype CD4 Pe Cy 5.5, CD8 APC Cy7, CD27 APC, CD45RA PE; B-cell phenotype CD19 PE, CD27 APC, IgD PE Cy5.5 (eBioscience, CA, USA), and ACE CD143 FITC (R&D Systems, Inc, Minneapolis, USA). After 30 minutes of incubation with monoclonal antibodies, in the dark and at 4°C, the cells were washed with PBS and centrifuged. Living cells (based on forward and side scatter) were acquired in the BD FACSCanto™ II using the DIVA software (Becton Dickinson, USA).

For the metabolic parameters, serum of studied individuals was previously isolated by centrifugation and frozen stored until the use. Measurement of metabolic parameters was performed in the laboratory central - Hospital São Paulo/UNIFESP.

## Statistics

Data are presented as mean ± standard deviation (SD). To test the normality of data it was used the Shapiro-Wilk test. We used p-value for interindividual differences in each variable since individuals age differently (biological aging) and thus, physiological parameters could be affected by genetics, lifestyle, nutrition, and comorbidities. P<0.05 was considered significant.

## Results

Table 1 shows that older adults are heterogeneous for some physiological parameters such as glucose, urea, Glycated hemoglobin (Hbglic), and C-reactive protein (CRP).

**Table 1.** Physiological parameters observed in older adults

|  | Metabolic parameters |  |  |  |  |  |  |  |  |
| --- | --- | --- | --- | --- | --- | --- | --- | --- | --- |
|  | Cholesterol<br>mg/dL | LDL<br>mg/dL | Triglycerides<br>mg/dL | Glucose<br>mg/dL | Urea<br>mg/dL | Creatinine<br>mg/dL | Albumin<br>mg/dL | HbGlic<br>mg/dL | CRP<br>mg/dL |
|  | 207 | 137 | 152 | 80 | 30 | 0.86 | 3.8 | 5.9 | 7.3 |
|  | 253 | 176 | 152 | 86 | 40 | 0.73 | 4.1 | 6.2 | 4.1 |
|  | 181 | 96 | 130 | 137 | 28 | 0.84 | 3.2 | 7.9 | 6.0 |
|  | 223 | 150 | 149 | 83 | 28 | 0.68 | 4.2 | 5.5 | 23.1 |
|  | 249 | 186 | 163 | 89 | 29 | 0.79 | 3.8 | 5.8 | 4.6 |
|  | 191 | 125 | 130 | 165 | 28 | 1.01 | 3.4 | 6.0 | 0.6 |
| Mean | 217.3 | 145 | 146 | 106.7 | 30.5 | 0.82 | 3.8 | 6.2 | 7.6 |
| ± SD | 27.2 | 30.4 | 12.1 | 32.5 | 4.3 | 0.1 | 0.4 | 0.8 | 7.2 |
| p | >0.100 | >0.100 | >0.100 | 0.047 | 0.017 | >0.100 | >0.100 | 0.022 | 0.026 |

Table 2 and figures 2, 3, and 4 show that CD143 (ACE1) is expressed in almost 100% of lymphocytes, whereas is is expressed in 56.9% (mean) of non-lymphocytes. CD8^+^ T cells presented the highest expression (98.4%), followed by CD19^+^ B cells (93.7%) and CD4^+^ T cells (90.7%). In T cells, ACE1 is expressed in all phenotypes (naive, central memory, effector memory, and effector memory re-expressing CD45RA - EMRA). In B cells, ACE was expressed in naive, unswitched memory, switched memory and double negative cells.

**Table 2.** CD143 (ACE1) expression in lymphocytes and non-lymphocytes

|  | Lymphocytes |  |  | Non-lymphocytes% |
| --- | --- | --- | --- | --- |
|  | CD4<sup>+</sup>CD143<sup>+</sup><br>% | CD8<sup>+</sup>CD143<sup>+</sup><br>% | CD19<sup>+</sup>CD143<sup>+</sup><br>% |  |
|  | 84.8 | 97.1 | 90.5 | 74.6 |
|  | 77.6 | 96.7 | 90.6 | 35.4 |
|  | 96.9 | 99.0 | 91.4 | 47.7 |
|  | 98.8 | 99.6 | 99.0 | 75.0 |
|  | 87.8 | 98.5 | 95.7 | 32.9 |
|  | 98.3 | 99.6 | 94.9 | 75.9 |
| Mean | 90.7 | 98.4 | 93.7 | 56.9 |
| ± SD | 8.7 | 1.3 | 3.4 | 20.6 |
| p | >0.150 | >0.150 | >0.150 | 0.081 |

**Figure 2.**
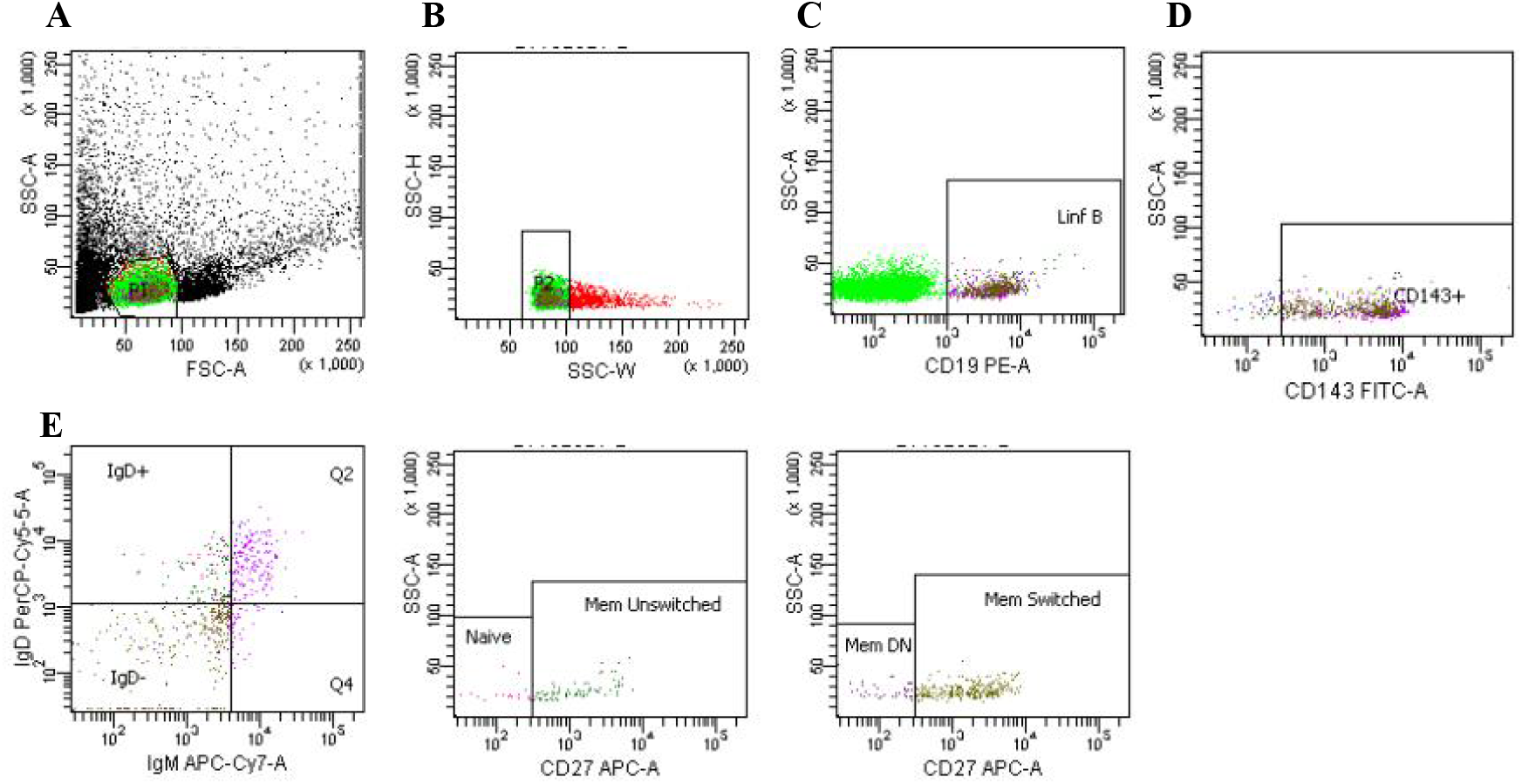
Flow cytometry gating strategy for B cell phenotypes and CD143 expression A) All cells and gate for lymphocyte (green) based on forward scatter (FSC-A) and side scatter (SSC-A) B) Doublets exclusion (from the lymphocyte gate) C) CD19^+^ B cells (from the doublets exclusion gate) D)CD143^+^ACE1 cells (from the CD19^+^ B cells gate) E)B cells phenotypes and CD143^+^ - IgM^+^IgD^+^CD27^-^ (naive); IgM^low^IgD^-^CD27^+^ (memory unswitched - Mem unswitched); IgM^-^IgD^-^CD27^+^ (memory switched - Mem switched); IgM^+^IgD^-^CD27^-^ (memory double negative - Mem DN)

**Figure 3.**
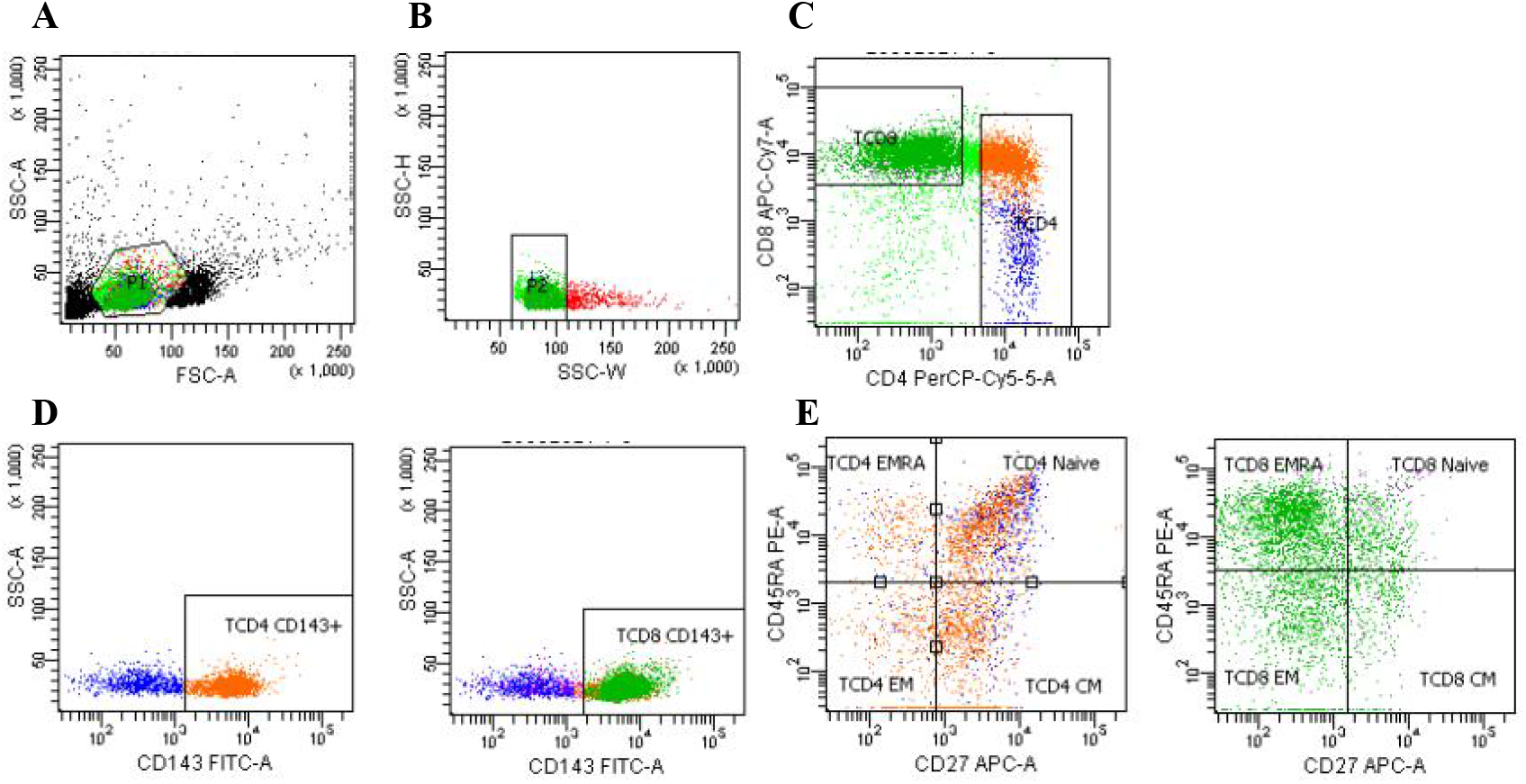
Flow cytometry gating strategy for T cell phenotypes and CD143 expression A) All cells and gate for lymphocyte (green) based on forward scatter (FSC-A) and side scatter (SSC-A) B) Doublets exclusion (from the lymphocyte gate) C) CD4^+^ and CD8^+^ T cells (from the doublets exclusion gate) D)CD143^+^ACE1 cells (from the CD4^+^ and CD8^+^ T cells gate) E)T cells phenotypes and CD143^+^ : CD45RA^+^CD27^-^ (naive); CD45RA^-^CD27^+^ (central memory - CM); CD45RA^-^CD27^-^ (effector memory - EM); CD45RA^+^CD27^-^ (effector memory re-expressing CD45RA - EMRA)

**Figure 4.**
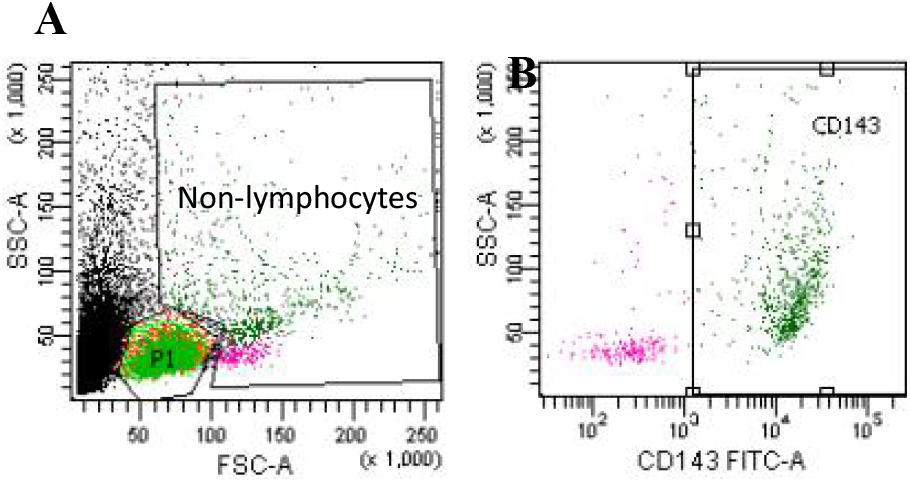
Flow cytometry gating strategy for non-lymphocytes and CD143 expression A) All cells and gate for lymphocyte (P1) and non-lymphocyte based on forward scatter (FSC-A) and side scatter (SSC-A) B) CD143^+^ ACE1 cells (from the non-lymphocyte gate)

Table 3 shows that both male and female present characteristics of senescent T cells such as low expression of Naive CD4^+^ and CD8^+^ T cells and high expression of EMRA CD8^+^ T cells.

**Table 3.**
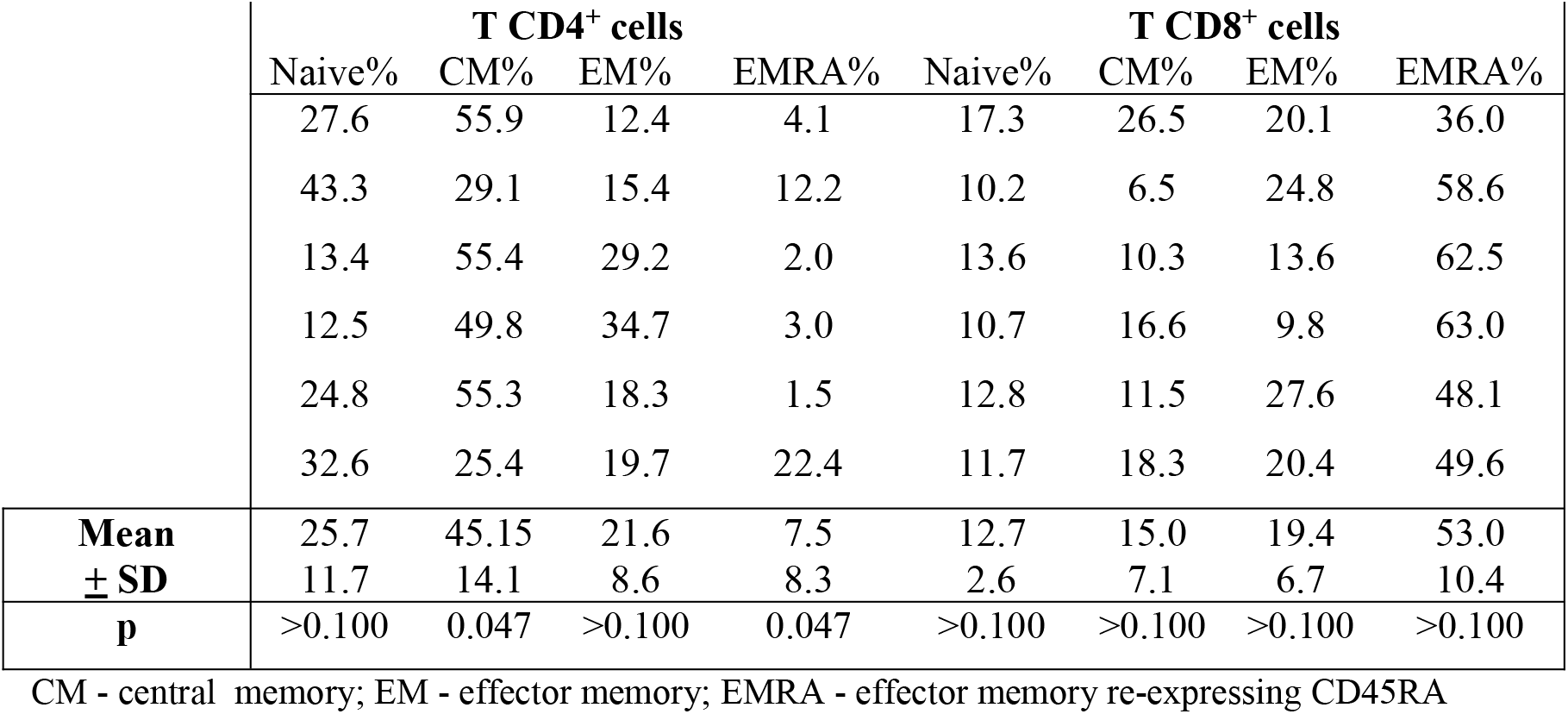
Phenotypes of CD4+ and CD8+ T cells

Table 4 shows that those aging adults with lower percentages of Naive B cells also presented a higher percentage of double negative (DN) memory B cells

**Table 4.** Phenotypes of CD19^+^ cells

|  | CD19<sup>+</sup> cells |  |  |  |
| --- | --- | --- | --- | --- |
|  | Naive % | Uns Memory % | S Memory % | DN Memory % |
|  | 73.8 | 6.3 | 4.0 | 15.9 |
|  | 61.3 | 6.9 | 5.7 | 26.1 |
|  | 28.6 | 4.1 | 31.4 | 35.8 |
|  | 51.8 | 10.0 | 22.1 | 16.1 |
|  | 35.9 | 7.9 | 18.5 | 37.7 |
|  | 67.7 | 3.5 | 9.8 | 19.0 |
| **Mean** | 53.2 | 6.5 | 15.3 | 25.1 |
| **± SD** | 17.9 | 2.4 | 10.6 | 9.8 |
| **p** | >0.100 | >0.100 | >0.100 | >0.100 |

Uns Memory - unswitched memory; S Memory - switched memory; DN Memory - double negative memory

## Discussion

Our results show that for the studied population, chronological aging and biological aging don’t go at the same pace. Even individuals having a small chronological difference (64 to 67 years old), they are heterogeneous for physiological parameters such as glucose, urea, glycated hemoglobin (Hbglic), and C-reactive protein (CRP). Changes in the same functional parameters have been reported by Carlsson et al. [22] and Helmerson-Karlqvist [23] in healthy older adults. Carlsson’s study [22] found that CRP value was 2.6 with a coefficient variation of 1.4% whereas in our study, it was observed higher values of CRP in 5 out of 6 individuals. Increased CRP levels has been associated with inflammaging and our findings show that the studied population has changes in functional parameters which are likely associated with an inflammatory profile. [24]

The link between RAS and inflammation has been suggested but its role is not completely clear under physiological and pathological conditions. [25, 26] In addition, the association between ACE1 altered expression in tissues (brain, muscle, heart and vessels) and the development and progression of age-related conditions such as Alzheimer’s, sarcopenia, and cardiovascular disease has been suggested but results are controversial. [17, 27, 28, 29, 30]

There are few studies showing the association between ACE1 expression in cells from the immune system (monocytes, T cells) and the progression of kidney and cardiovascular disease. [9, 8, 31, 32]. Therefore, considering the lack of information on this issue, we questioned whether ACE1 (CD143) was highly expressed in cells from the immune system during the aging process. We found that ACE1 was expressed in almost 100% of T (CD4^+^, CD8^+^) and B lymphocytes and in all phenotypes of these cells. In non-lymphoid cells, ACE1 mean expression was 56,9%. In agreement with our findings, independent studies showed that T cells from healthy donors and monocytes from patients with congestive heart failure expressed ACE1, but there was no investigation on cell phenotype. [25, 26]. Our study is the first to show that either inexperienced (naive) or fully activated (memory) cells expresses ACE1. Our findings suggest that the expression of ACE1 in lymphoid and non-lymphoid cells reflects the health status since our studied population presented changes in physiological parameters and high levels of ACE1 expression on immune cells. Previous independent studies showed that patients with unstable angina [32] or acute myocardial infarction [33] presented higher expression of ACE1 in T cells and dendritic cells than controls subjects. In addition, markers of the cell (lymphoid and non-lymphoid) functional status such as inflammatory or growth factors production could be modulated by ACE inhibitors (ACEi). Accordingly, mononuclear leukocytes from healthy subjects incubated with endotoxin exhibited high levels of tissue factor activity which was reduced in the presence of captopril in a dose-dependent pattern. This result could be related to the antithrombotic effect of ACEi. [34]. In patients with congestive heart failure, immune cells cultured with LPS secreted high levels of the pro-inflammatory TNF-alpha and these levels were significantly reduced in the presence of captopril. [35]

It can be proposed that mechanistically, AngII is produced by mononuclear cells/lymphocytes and at the same time AngII induces immunologic activation on these cells. Therefore, the inflammatory axis ACE1/AngII/AT_1_R and the counter-regulator ACE2/Ang1-7/MasR axis [36, 37] could play a role in chronic diseases, inflammaging and immunosenescence observed in older adults. Our studied population presented changes in some physiological parameters and increased levels of C-reactive protein (CRP). This inflammatory profile [24] in addition to more than 90% of T and B cells expressing ACE1 in our older adults suggest a correlation between aging, inflammaging and ACE1. Independent of the chronological age, inflammation (even if related to sub-clinical diseases) may be a contributor for disease progression when the balance with anti-inflammation is shifted. [38] In this context, the control of ACE1/ACE2 expression could be explored as a target for the balance of exacerbated inflammatory reactions. Considering that the equilibrium between ACE1 and ACE2 expression could play an important role for the healthy aging, our next studies will be focused on ACE1 and ACE2 expression in cells from the immune system.

The phenotype of T and B lymphocytes has been used to identify the senescence in immune cells. CD4^+^ T cells present changes during the aging process with decrease of Naive and increase of Effector Memory (EM) phenotypes whereas CD8^+^ T cells show decrease in Naive and increase in EM and Effector Memory re-expressing CD45RA (EMRA). [12, 39, 40] It has been shown the reduction of Naive B cells and no change for memory unswitched and memory switched B cells. In addition, it has been observed an increase in the percentage of double negative B cells. [41, 42, 43, 44] Using these phenotypes, we found similar senescent phenotype in some of the studied aging adults. The reduction of Naive lymphocytes has been related to impaired antigen responsiveness and for B cells it is observed decrease in the production of antibodies. [45, 46] The increased percentage of double-negative memory (DN memory) B cells has been linked to autoimmune diseases. [47, 48]. We found ACE1 expression in more than 90% of T cells and B cells and in all phenotypes. ACE1 was expressed in non-lymphocytes in a range of 32.9% to 75.9%. Our findings suggest that ACE1 could play a role in several processes linked to aging including the generation and activation of autoimmune cells, due to the experimental evidence that inhibitors of ACE suppress the autoimmune process in a number of autoimmune diseases such as EAE, arthritis, autoimmune myocarditis. [49]

The present study is the first to compare the expression of the protein ACE1 among different cell types, both lymphoid (CD4^+^ and CD8^+^ T cells, B cells), and non-lymphocytes in older adults. It was also observed that even though the individuals studied were in the early stage of chronological aging (64 to 67 years old), they presented heterogeneity in the physiological parameters, signs of inflammaging (increased CRP) and immunosenescence such as low expression of naive T and B cells in addition to the accumulation of terminally differentiated T CD8^+^ and double negative (DN) B cells. This study have limitations such as the small sample size and the lack of young adults for comparison. As an example, the subject with the highest CRP and albumin also exhibited a high percentage of ACE1 expression on T (CD4^+^, CD8^+^), B and non-lymphoid cells in addition to the lowest percentage of CD4^+^ naive cells, and the highest percentage of CD8^+^ terminally differentiated (EMRA) and DN B cells. However, due to the small sample size it was not possible to associate the high expression of ACE1 on immune cells with inflammaging and immunosenescence. It would bring important information to correlate physiological parameters/health status with ACE1 expression and to find out whether age and associated chronic diseases could lead to increased ACE1 expression. Moreover, we only have CRP as a marker of inflammaging and IL-6 and TNF-alpha would be desirable to complete our panel. Functional analysis are needed to clarify the impact of ACE1 expression on immune cells and whether ACE inhibitors (ACEi) and angiotensin receptor blockers (ARBs) administered to hypertensive patients affect somehow the immunity. Recently it was shown that the membrane-bound ACE2 acts as a receptor for SARS-CoV-2 but it is still under investigation the possible effects on RAS components (AngII, Ang1-7, ACE1, ACE2, AT1 and Mas) and whether ACEi and ARBs interfere with the mitigation of COVID-19. [50, 51, 52, 53, 54] Therefore, it is important to emphasize the negative impact of chronic diseases for the outcome of older adults during a viral infection and how ACE1/ACE2 expression in immune cells could bring information to diagnosis and treatment.

## Data Availability

All data produced in the present study are available upon reasonable request to the authors

## Acknowledgments

Pedro Destro has a CNPq fellowship, CAPES PrInt UNIFESP n° 88881.310735/2018-01

